# Development of a cloud framework for training and deployment of deep learning models in Radiology: automatic segmentation of the human spine from CT-scans as a case-study

**DOI:** 10.1101/2024.08.27.24312635

**Authors:** Rui Santos, Nicholas Bünger, Benedikt Herzog, Sebastiano Caprara

## Abstract

Advancements in artificial intelligence (AI) and the digitalization of healthcare are revolutionizing clinical practices, with the deployment of AI models playing a crucial role in enhancing diagnostic accuracy and treatment outcomes. Our current study aims at bridging image data collected in a clinical setting, with deployment of deep learning algorithms for the segmentation of the human spine. The developed pipeline takes a decentralized approach, where selected clinical images are sent to a trusted research environment, part of private tenant in a cloud service provider. As a use-case scenario, we used the TotalSegmentator CT-scan dataset, along with its annotated ground-truth spine data, to train a ResSegNet model native to the MONAI-Label framework. Training and validation were conducted using high performance GPUs available on demand in the Trusted Research Environment. Segmentation model performance benchmarking involved metrics such as dice score, intersection over union, accuracy, precision, sensitivity, specificity, bounding F1 score, Cohen’s kappa, area under the curve, and Hausdorff distance. To further assess model robustness, we also trained a state-of-the-art nnU-Net model using the same dataset and compared both models with a pre-trained spine segmentation model available within MONAI-Label. The ResSegNet model, deployable via MONAI-Label, demonstrated performance comparable to the state-of-the-art nnU-Net framework, with both models showing strong results across multiple segmentation metrics. This study successfully trained, evaluated and deployed a decentralized deep learning model for CT-scan spine segmentation in a cloud environment. This new model was validated against state-of-the-art alternatives. This comprehensive comparison highlights the value of the MONAI-Label as an effective tool for label generation, model training, and deployment, further highlighting its user-friendly nature and ease of deployment in clinical and research settings. Further we also demonstrate that such tools can be deployed in private and safe decentralized cloud environments for clinical use.

**Author Summary:** In the rapidly evolving field of medical imaging, the integration of artificial intelligence (AI) and cloud computing is becoming increasingly critical for advancing diagnostic and treatment capabilities. To address the growing demand for flexible digital frameworks in the clinical environment supporting the deployment of data-driven applications, we have developed and deployed a cloud-based Trusted Research Environment designed specifically for training, validation, and deployment of deep learning models focused on semantic segmentation in musculoskeletal radiology. This environment facilitates the efficient handling of large datasets and the accessibility of algorithmic output for the physicians, optimizing the interface between development and clinical translation. The established framework enables significant improvements in the deployment of deep learning tools for image analysis in the clinical setting. In our current use-case, we have utilized this environment to train and evaluate two advanced deep learning models for the segmentation of the human spine from CT scans. By leveraging the computational power and flexibility of the cloud-based infrastructure, we were able to perform rigorous training and comparison of these models, aiming to enhance the accuracy and reliability of spine segmentation in clinical practice. This approach not only streamlines the process of model development but also provides valuable insights into the performance and potential clinical applications of these AI-driven segmentation tools.

## Introduction

With the inflection point of Artificial Intelligence (AI) algorithms, with its deep learning (DL) models and frameworks (1), medical operations are facing the paradigm shift of both digitalization and DL models deployment. Out of all clinical specialties, Radiology is poised to be at the forefront of this transformation due to its inherent reliance on visual data and the well-established role of image analysis in diagnosis and treatment planning (2,3). As such, there is a strong research focus on developing and deploying AI models within the Radiology field. As an example, in the past 5 years (2019-2023), a total of 11’392 research manuscripts (PubMed search) were related in some form with the thematic of radiology and deep learning (DL).

Indeed, medical imaging is a key area for the development of high added-value DL models that streamline and accelerate the process of image segmentation and classification, which is a major time-consuming task for physicians (4).

Several AI tools have been developed with the objective of accelerating medical image segmentation. Recently, the development of the TotalSegmentator DL model (5), trained to segment 117 anatomical structures (version 2.0), from CT datasets, demonstrates the immense possibilities that DL-powered medical image segmentation offers. For the development of such a powerful DL segmentation model, the nnU-Net framework was used (6). This framework allows the training of state-of-the art segmentations models, based on the U-Net DL architecture (7), with minimal to no coding needed. Still, these tools are limited in their deployment in daily clinical scenarios. As an easier and user-oriented approach, the open-source framework MONAI (Medical Open Network for Artificial Intelligence) (8) has been developed for the training and easier deployment of automatic medical image segmentation models. At the forefront of MONAI, the sub-framework MONAI-Label (9) connects DL architectures and pretrained models with open-source graphic user interfaces (GUI) such as 3D Slicer and OHIF (Open Health Imaging Foundation Viewer), enabling active learning with direct visualization and correction of segmentation results (10,11).

As optimal environment for the deployment of such aforementioned frameworks, the adoption of cloud computing in medical research setting, offers many benefits: it fosters improved collaboration among physicians and researchers, provides a decentralized and scalable computing infrastructure and greater accessibility to tools and services such as giving support for data annotation. Furthermore, cloud-based infrastructure offers simplified management of resources in a cost-effective manner, allowing medical research centers to focus resources on data analysis and clinical relevance of research questions rather than IT maintenance (12,13).

In this study, we have developed a cloud computing framework leveraging the open-source pipeline of MONAI-Label for the development and deployment for clinical research teams of an interactive spine segmentation model out of CT data. Moreover, we benchmarked the model output with an in-house trained nnU-Net model, both trained and validated with the same collection of images (TotalSegmentator open-source dataset). We also compare it with a pretrained model for spine segmentation present in MONAI-Label, trained on the VERSE dataset (14). We demonstrate that the MONAI-Label developed model produces similar segmentation results to the model trained with the nnU-Net framework yet providing a far simpler vertical integration in a clinical research setting. This *de novo* trained model also considerably outperforms the pretrained spine segmentation model present in MONAI-Label *ab initio*. Moreover, the presented research highlights the use of MONAI-Label as a powerful tool for developing DL models and automatically generating anatomical segmentation of anatomical structures which may support surgical planning, diagnostics and future creation of digital twins of patients (15,16).

## Materials and Methods

### Architecture of a trusted cloud research environment for deep learning model training and deployment

The framework introduced in this work is based on a private cloud environment composed of private endpoints, i.e., a pointer to the location of the data, which allows the hospital network to be extended in a private cloud space (figure 1A). Through an Application Programming Interface (API) Manager, the DICOM (Digital Imaging and Communications in Medicine standard) or NIFTI (Neuroimaging Informatics Technology Initiative) storage locations connect with cloud computational nodes (with adequate hardware accelerators, such as graphic processing units – GPUs), and a centralized management dashboard that allows the definition of strict governance policies, as the ones in place at healthcare institutions, granting secure and private access to specified users. Through the combination of the cloud services mentioned above, the framework composed of 3D Slicer and MONAI Label-based computational nodes was developed. In this way, scalable computation resources are connected with a Graphical User Interface (GUI) thanks to the open-source tool 3D Slicer, as shown on figure 1b.

**Figure 1:**
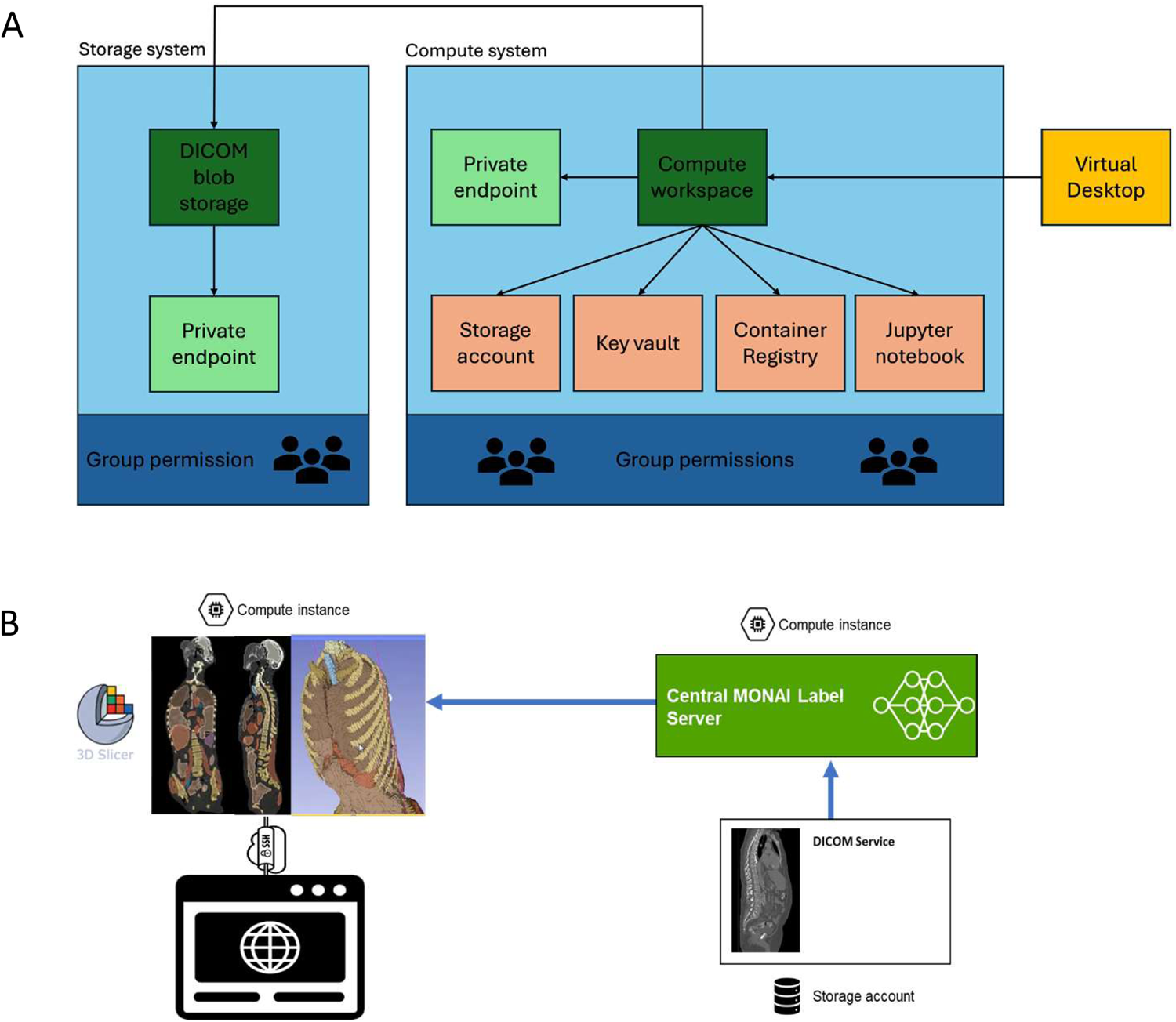
Architecture of the cloud infrastructure behind the trusted research environment and deployed framework with 3D Slicer and MONAI-Label. **(A)** Overview of the components of the architecture of the trusted research environment: storage and compute. The first one has a DICOM blob storage which is exposed with a private endpoint; while the second one has more components. The compute workspace is exposed as well with a private endpoint, while the other component such as storage account, key vault Jupyter notebook and container registry are connected to the workspace. Both systems have group permissions that can be managed individually. To access the trusted research environment, users can use a Virtual Desktop application (B) Using two compute nodes, 3D Slicer and MONAI Label are deployed separately. With an SSH connection users can access an online environment to use the 3D Slicer interface to interact with the model’s output that runs on the first compute node (from the left side), which process the DICOM images retrieved by the second compute node on which MONAI Label is running from the DICOM blob storage.

For segmentation model training in the trusted research environment, the TotalSegmentator open-source image dataset was stored in a project-specific NIFTI storage and connected with computational resources embedded in the computational node via the API Manager.

For segmentation inference, an additional API allows the access to DICOM images stored in the clinical PACS (Picture Archiving and Communication System), retrieve these images to a DICOM storage location in the cloud which then directly connects to the MONAI Label computational node for automatic segmentation processing. The connection to the GUI results in a user-friendly environment where radiologists and other physicians can visualize the segmentation results, correct them, and potentially fine-tuning the models by sending the corrected results back to MONAI Label central instance to update the training.

### Resources for deep learning model training

The TotalSegmentator dataset composed of CT images and their ground-truth vertebrae and sacrum segmentations (part of the open-source dataset) was used for model training in either the MONAI-Label or nnU-Net frameworks (5) (https://zenodo.org/records/10047292, with an average isotropic pixel size of 1.5 mm). The dataset was divided into training and inference dataset. 964 images were randomly chosen to be part of the training dataset, while the remaining 108 were used for validation (the open-source version of the dataset contains more than 1200 images, nonetheless, some cannot be used with the current version, 2.0.1). The dataset was preprocessed in accordance with the requirements of both MONAI-Label and nnU-Net (instructions found at https://github.com/Project-MONAI/MONAILabel?tab=readme-ov-file#getting-started-with-monai-label and https://github.com/MIC-DKFZ/nnUNet). The dataset was uploaded into a storage location in the developed cloud-based Trusted Research Environment. The default segmentation task in MONAI-Label, based on a ResSegNet architecture (17), was used for spine segmentation model training. The training was performed in 4 A100 GPU computational nodes, for 1000 epochs, 241 iterations each, taking *circa* 25 hours of compute time. Since for both training and results analysis a graphic user interface is necessary, the open-source 3D Slicer tool was used in a container (https://github.com/pieper/SlicerDockers, v5.0.3). No validation split was used in this training as model benchmarking would be done with an already assigned dataset (remaining 108 images available). Default segmentation pipeline scripts were used with no modifications (segmentation.py, part of the radiology app from the MONAI-Label framework with a tutorial found here: https://github.com/Project-MONAI/tutorials/tree/main/monailabel). The nnU-Net model training was performed on one A100 compute node for 1000 epochs, 250 iterations each, over the course of 15 hours of training. No validation split was used (fold 0), as model benchmarking would be done with an already assigned dataset (same 108 datasets used to validate the MONAI-Label model). Default nnU-Net pipeline scripts were used with no modifications, following the developers tutorial.

### Deep learning model benchmarking

108 CT scans of the TotalSegmentator dataset (not part of the dataset for DL model training) were used for model validation. We calculated different segmentation metrics to benchmark both deep learning models’ segmentation results such as dice score, intersection over union, accuracy, precision, sensitivity, specificity, bounding F1 score, recall, area under the curve (AUC) and the Hausdorff distance between segmentation predictions and their ground truth (18). Each metric was calculated with custom scripts (available upon request). Calculation of distances between ground truth and predicted segmentations was done in the open-source software Meshlab (https://www.meshlab.net/) v2022.02 with built-in functions. P-values were calculated based on a non-paired t-student function.

## Results

### Benefits of a decentralized cloud infrastructure for DL model training and deployment

Deploying a decentralized trusted research environment on the cloud for training and deployment of deep learning models present two major benefits: on one hand the scalability and flexibility to compose a tailored DL model for a specific research or clinical question, on the other hand, the possibility to deploy the developed models in clinical workflows using the same architecture.

Depending on the size and architecture of the model, the compute resources of the computational nodes can be quickly scaled-up or scaled-down, providing the creation of an optimized setup for each use-case requiring a training, validation and/or deployment of a research or clinical DL model. Further, this decentralized approach makes it far simpler to securely and privately connect different medical devices (CT scanners, MRI machines, ultrasound machines, etc.) as endpoints to the developed trusted research environment and get outputs in real-time.

Within the MONAI Label framework, multiple models can easily be retrieved and fine-tuned by labeling additional data via a user-friendly GUI, in this way a local version can be trained. The versatility of this process enables various healthcare operators, e.g. radiologists, to retrain models on top of open-source available models (transfer learning). Furthermore, they can easily test models that have been previously created in the framework and validate them thanks to commonly available software for 3D visualization. This way, the interaction between model’s development and radiologists is considerably improved and physicians are a central component in the development of new ML models.

### Training a ResSegNet based CT-scan spine segmentation model within the MONAI-Label framework: spineCT-ResSegNet model

Out of 1072 images present in the TotalSegmentator, 964 images (90% of the dataset), were randomly chosen to compose the training dataset, while 108 (remaining dataset) served as the validation dataset. Default 3D ResSegNet parameters of the segmentation (ResSegNet) task of MONAI-Label radiology app, were used. Qualitative analysis of the segmentation results (figure 2A) demonstrates the capacity of robust prediction of the entire human spine and sacrum (25 segmented classes). Vertebral bodies identification is correctly done, with their shape being in accordance with anatomical expectancy. Quantitative segmentation quality metrics over all the 25 classes reveal a robust model for spine segmentation from CT-scans with an average dice score of 0.856±0.066, an average intersection over union score of 0.801±0.074, average accuracy of 1.000±0.000, a mean precision of 0.862±0.066, a mean sensitivity (or recall) of 0.861±0.070, a mean specificity of 1.000±0.000, an average bounding F1 score of 0.837±0.057, a mean area under the curve (AUC) of 0.932±0.034 and a Kappa of 0.853±0.068 (figure 2B, all numbers rounded to 3 decimal places). Detailed results, with 95% confidence intervals, are found in supplementary figure 1.

**Figure 2.**
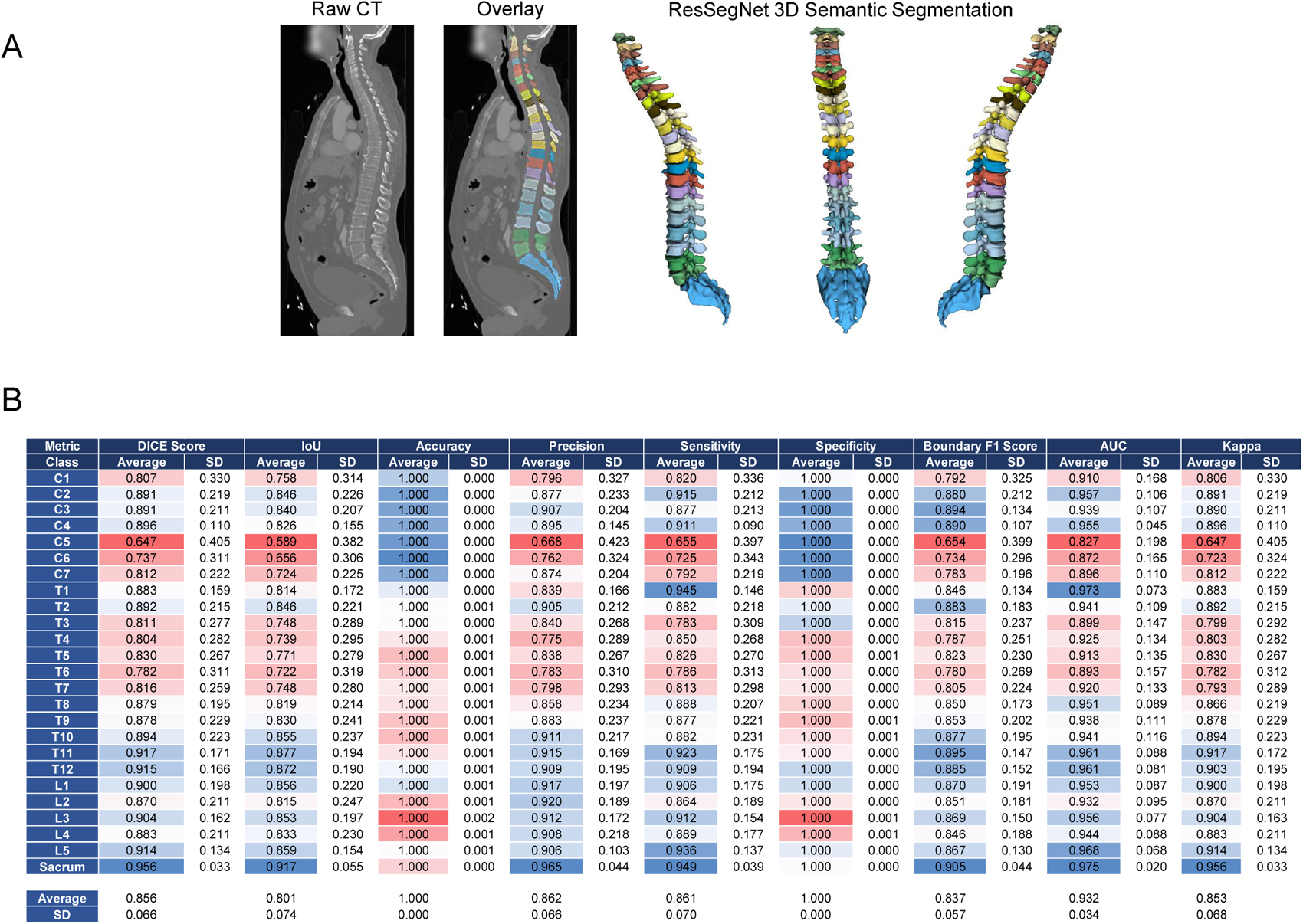
SpineCT-ResSegNet spine segmentation model predictions and segmentation metrics. (A) Example of segmentation results with the spineCT-ResSegNet, a trained MONAI-Label native model, in both 2D and 3D visualizations. Vertebrae are color coded. The entire human spine is represented in the image: cervical spine (C1-C7), thoracic spine (T1-T12), Lumbar spine (L1-L5) and sacrum. (B) Validation metrics for spineCT-ResSegNet spine segmentation model, based on the prediction of 108 cases. Color encodes the accuracy of segmentation class predictions. Blue indicates high similarity to the ground truth, while red represents increasing deviation from the actual values.

### Training a nnU-Net based CT-scan spine segmentation model: spineCT-nnUNet model

Serving as a benchmark for DL segmentation model comparison, we trained the nnU-Net framework on the same dataset 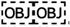, with the 3D-UNet full resolution model (1.5 mm), using the default configuration and trained the model over 1000 epochs. Validation was performed with the same 108 CT scans as for the MONAI-Label ResSegNet model, native to MONAI-Label. Segmentation output is qualitatively robust with anatomical structures, i.e. vertebrae, are localized to their correct position and have a correct tridimensional conformational (figure 3A). 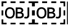 between the segmentation prediction and their ground-truth, reveal a robust prediction model, with an average dice score over the 25 classes (anatomical structures) of 0.914±0.036, an average intersection over union of 0.879±0.045, an accuracy of 1.000±0.000, a precision of 0.911±0.046, a sensitivity (or recall) of 0.910±0.050, a specificity of 1.000±0.000, a boundary F1 score of 0.908±0.029, an AUC score of 0.961±0.018 and a Kappa score of 0.904±0.049 (figure 3B, 5 all numbers rounded to 3 decimal places). Detailed results, with 95% confidence intervals, are found in supplementary figure 2.

**Figure 3.**
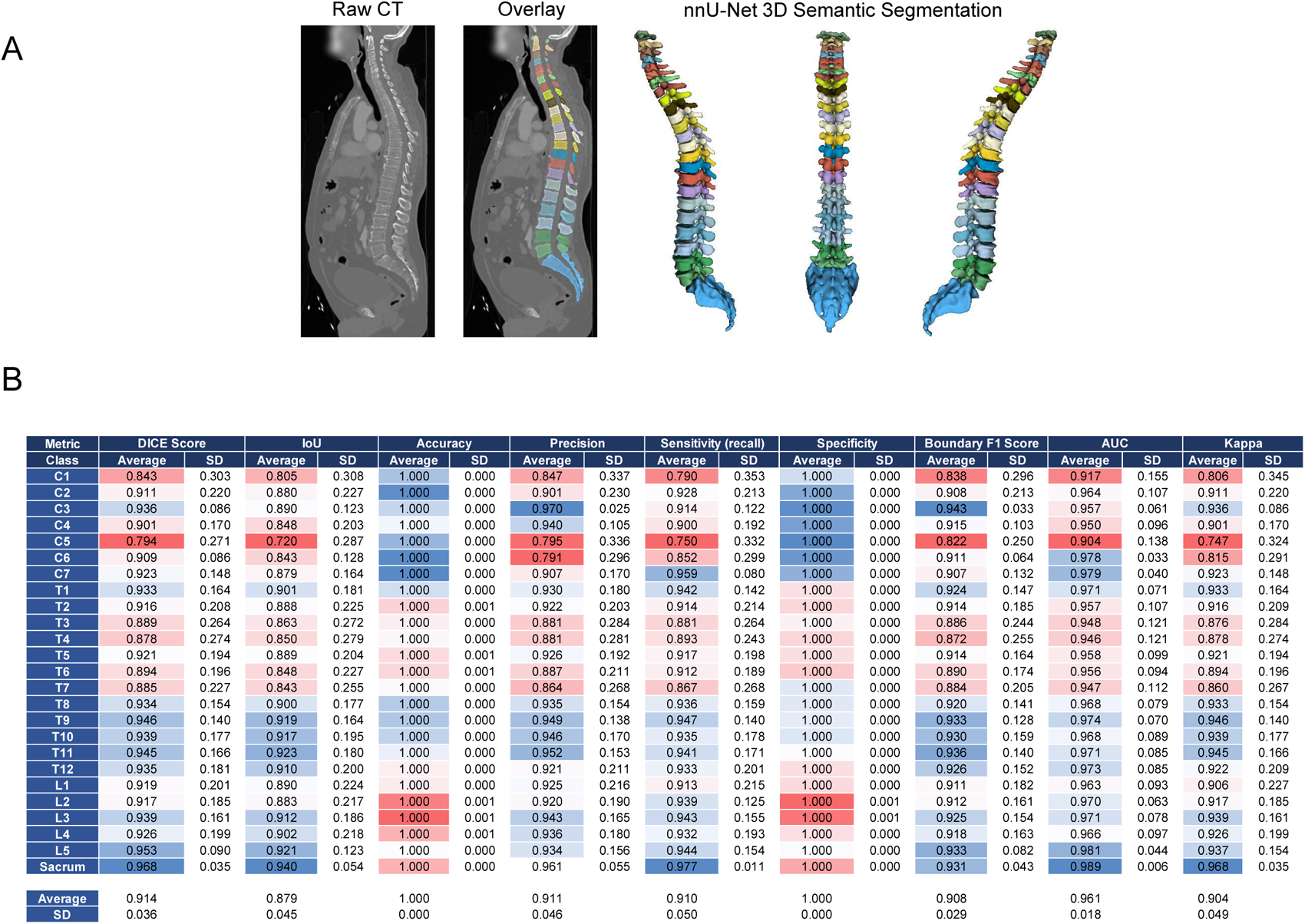
SpineCT-nnUNet spine segmentation model predictions and segmentation metrics. (A) Example of segmentation results with the nnU-Net framework derived model (3D-full resolution) in both 2D and 3D visualizations. Vertebrae are color coded. The entire human spine is represented in the image: cervical spine (C1-C7), thoracic spine (T1-T12), Lumbar spine (L1-L5) and sacrum. (B) Validation metrics for spineCT-nnUNet spine segmentation model. Color encodes the accuracy of segmentation class predictions. Blue indicates high similarity to the ground truth, while red represents increasing deviation from the actual values.

### Comparison of spineCT-ResSegNet, spineCT-nnUNet and pretrained spine segmentation models

To further compare the segmentation predictions between the ResSegNet model and the nnU-Net model to the respective ground truth, we overlaid selected examples as exemplified in figure 4A (spineCT-ResSegNet) and 4C (spineCT-nnUNet). Hausdorff distances between each segmentation model predictions and their ground truths were calculated (spineCT-ResSegNet and spineCT-nnUNet in figure 4B and figure 4D respectively). The spineCT-ResSegNet model presents an average Hausdorff distance of 2.604±0.450 mm while the spineCT-nnUNet model predictions present a Hausdorff distance of 2.214±0.398. While the nnU-Net based model outperforms the ResSegNet model in regard to the quality metric of Hausdorff distance, the same is not observed for the remaining metrics calculated. Thus, we find sufficient evidence to say both models produce on-par segmentation results (supplementary figure 5A, all p-valued rounded to 5 decimal plates).

**Figure 4.**
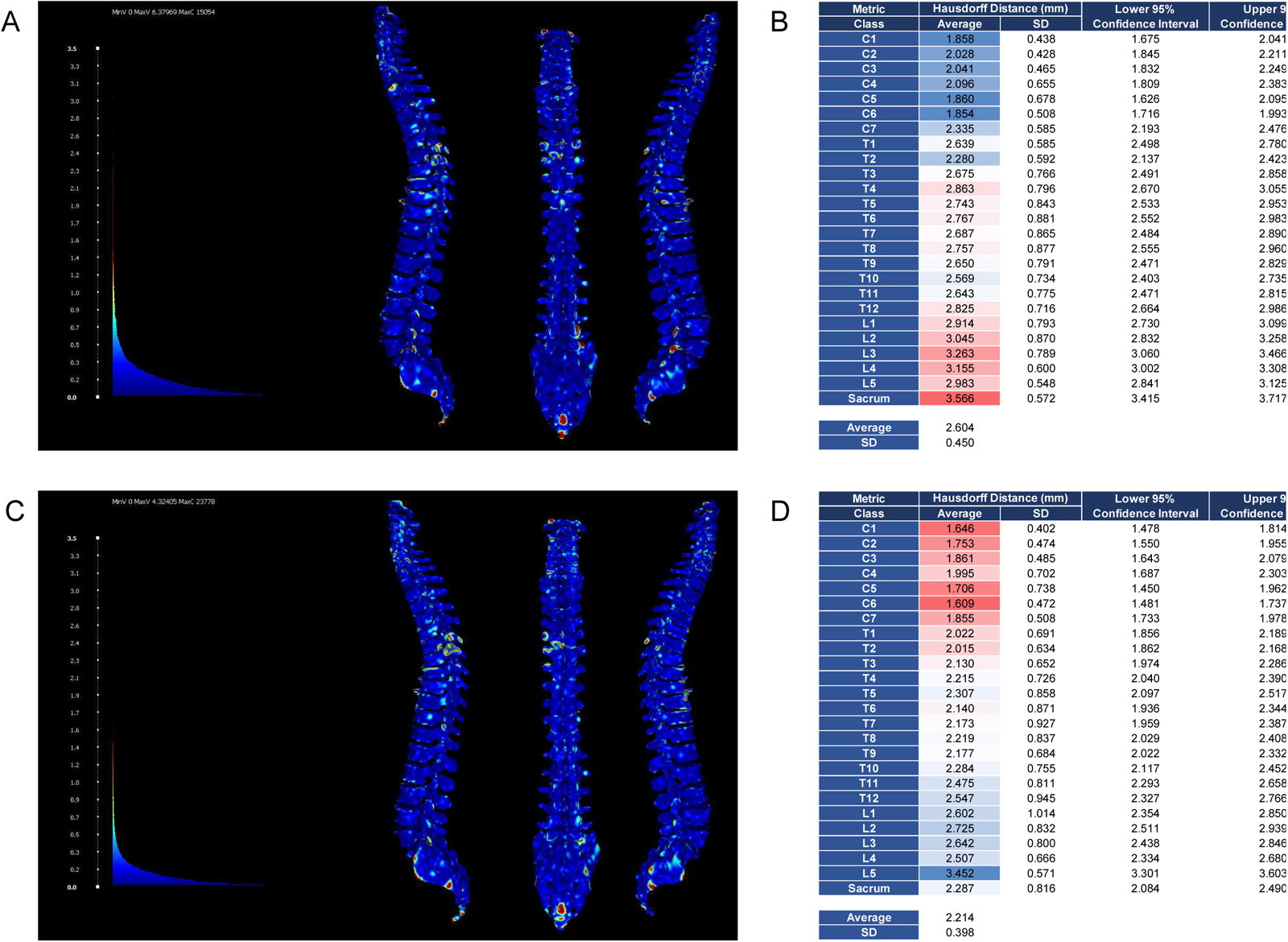
Comparison of spineCT-ResSegNet and spineCT-nnUNet semantic segmentation models. (A) Example of the Hausdorff distance calculation between the spineCT-ResSegNet model (native to MONAI-Label) predictions and ground truth. (B) Average Hausdorff distance calculation for spineCT-ResSegNet predictions versus their ground truth. Predictions with the spineCT-ResSegNet model average a Hausdorff Distance of 2.604±0.450 mm to the ground truth. (C) Example of the Hausdorff distance calculation between the spineCT-nnUNet model predictions and ground truth. (D) Average Hausdorff distance calculation for spineCT-nnUNet predictions versus their ground truth. Predictions with the spineCT-nnUNet model average a Hausdorff Distance of 2.214±0.398 mm to the ground truth. Color encoding in (B) and (D) correlates with the accuracy of segmentation class predictions. Blue indicates high similarity to the ground truth, while red represents increasing deviation from the actual values.

We also benchmarked the pretrained vertebrae segmentation model which is part of the MONAI-Label core. The model spine segmentation predictions average a dice score over the 24 classes (as this model only predicts from C1 to L5) of 0.601±0.153, an average intersection over union of 0.533±0.154, an accuracy of 1.000±0.000, a precision of 0.697±0.150, a sensitivity (or recall) of 0.565±0.157, a specificity of 1.000±0.000, a boundary F1 score of 0.588±0.144, an AUC score of 0.785±0.080 and a Kappa score of 0.597±0.151 (supplementary figure 3, all numbers rounded to 3 decimal places). The output segmentations present a Hausdorff distance of 3.129±0.322 mm on average. Thus, we can demonstrate that the *de novo* trained model spineCT-ResSegNet outperforms the pretrained model in MONAI-Label in most quality metrics calculated (supplementary figure 5B, all p-valued rounded to 5 decimal plates). Furthermore, it also trained in one more class, the sacrum.

## Discussion

A decentralized trusted research environment as a core of novel digital ecosystems in medical research has the potential to simplify communication between physicians and researchers, enable effective testing and validation of novel tools, provide secure and private data exchange channels - and ultimately reduce costs by sharing computational resources, with non-complicated scalability. Further benefits are the ability to access common datasets, shared code, and validation methods at any time reduces errors and improves usability - and thus accelerates the clinical translation of digital technologies. Altogether, such platforms facilitate testing and validation of digital and DL pipelines, making data-driven medical research more reproducible and better suited for the deployment in clinical settings. With these principles in mind, we developed a radiology-specific cloud infrastructure, based on a combination of DICOM storage services and MONAI-Label-compatible computational nodes for the training and deployment of deep learning segmentation models.

As a case-study, we trained and deployed a ResSegNet model for human spine segmentation out the open-source dataset of CT-scans from TotalSegmentator. This model was trained as part of the MONAI-Label (8,19) framework. As a benchmark, we used the nnU-Net framework (6) and also trained a segmentation model with the same training and inference dataset. Both models yielded similar quality metric results with only average Hausdorff distance being noticeable better for the nnU-Net model, than for the ResSegNet model (figure 4 and supplementary figure 5). Nonetheless, given the native deployment of the ResSegNet model via MONAI-Label, we can directly deploy it via a graphic user interface such as 3D Slicer or OHIF (10,11), which is not possible with the nnU-Net, at least not in a straightforward way. This in-place visualization provides direct access to the segmentation results to the health professionals who can use it to make informed decisions, e.g. for surgical planning.

We also compared our trained model, spineCT-ResSegNet, with the pre-trained and pre-built spine segmentation model which MONAI-Label already offers (14). Our newly trailed model greatly outperforms the prebuilt model in all metrics, providing substantially more accurate segmentations (supplemental figure 4 and 5).

While our developed models underperformed compared with the dice scores reported for the TotalSegmentator model (average 0.95 for vertebrae) (5), this nnU-Net based model was trained for 8000 epochs compared to ours which was trained for only 1000 epochs. This meant a far lower training time for our model, fewer computational resources were needed, while still performing adequately for the task at hand.

Naturally, given the dice coefficient of around 0.85 and a hausdorff coefficient of approximately 2.6 mm, the ResSegNet model incurs in mispredictions and proper segmentation correction may be necessary. Nonetheless, the iterative nature and easy of use of MONAI-Label and a chosen GUI such as 3d Slicer, makes any necessary corrections easy to perform. Further, this corrections may be included as part of a subsequent finetuned model, as part of the labelling correction process, which will then, increase the predictive power of the model.

## Conclusion

The present research demonstrates the potential of cloud-based deep learning for training and deployment of medical image segmentation models, in a trusted research environment. By leveraging the efficiency and scalability of cloud computing, we have created a framework that empowers medical professionals, such as radiologists, with faster, more accessible segmentation tools, based on the MONAI-Label open-source framework. This paves the way for improved diagnostics, treatment planning, and ultimately, better patient outcomes.

## Data Availability

The data used in this study is publicly available at https://github.com/wasserth/TotalSegmentator and https://zenodo.org/records/10047292

https://zenodo.org/records/10047292

## Abbreviations

DL: deep learning
AI: artificial intelligence
AUC: area under the curve

## Author Contribution

NB developed and deployed the trusted cloud environment. RS worked on the development and testing of the deep learning segmentation models. BH assisted NB in the infrastructure setting of the trusted cloud environment. NB, RS and SC wrote the manuscript and designed the figures. SC supervised the design of the infrastructure and pipeline and contributed to the design of the research and interpretation of the results. MF contributed to the final version of the manuscript.

## Acknowledgements

The authors acknowledge the Digital Society Initiative, University of Zurich. This work is part of the DSI-Infrastructure / Lab-Program.

---

**Supplemental Figure 1.**
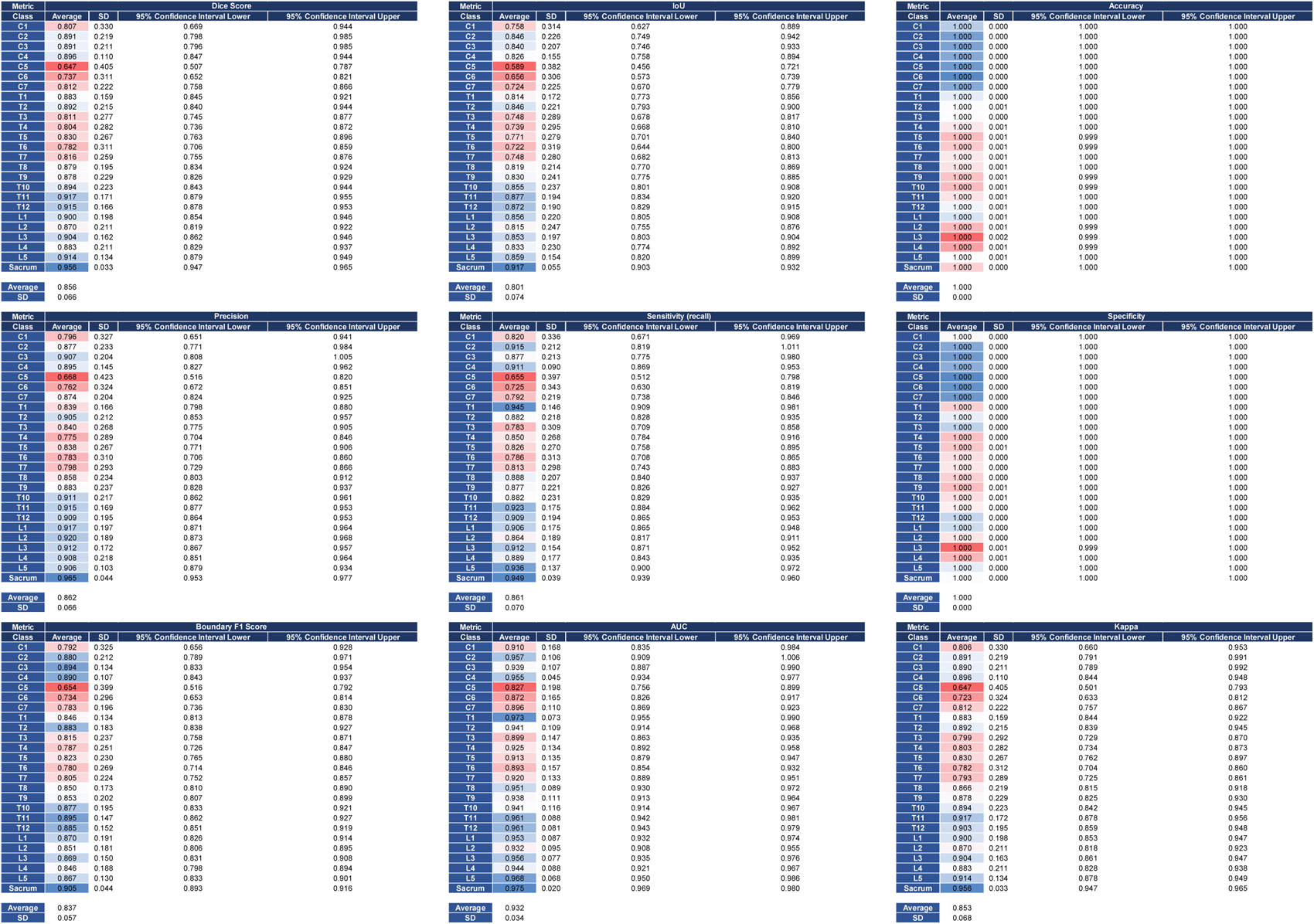
Comprehensive metrics evaluation of the spineCT-ResSegNet. Average metrics for the spineCT-ResSegNet model based on comparison between the predictions of the model on 108 datasets and their ground truth.

**Supplemental Figure 2.**
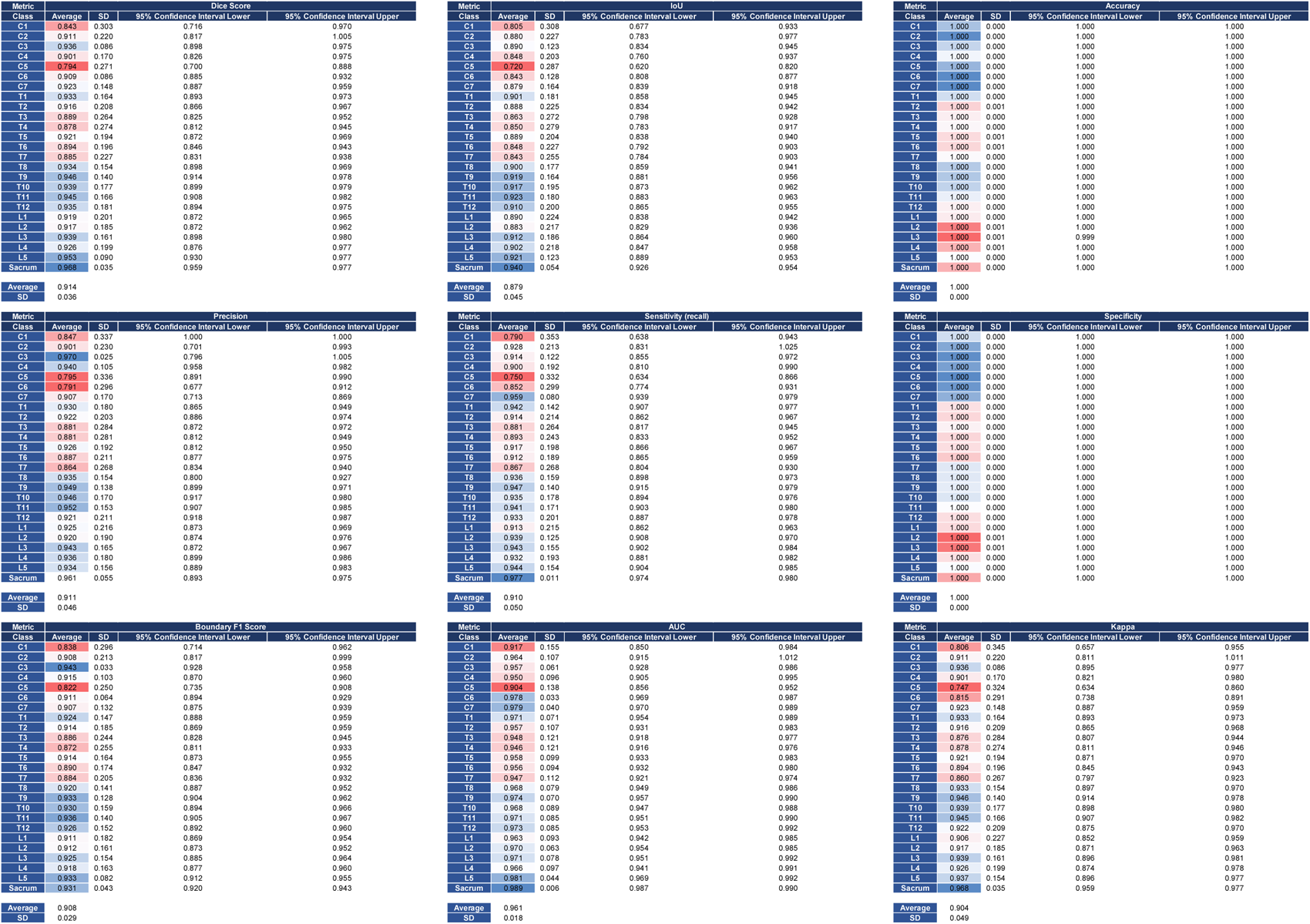
Comprehensive metrics evaluation of the spineCT-nnUNet. Average metrics for the spineCT-ResSegNet model based on comparison between the predictions of the model on 108 datasets and their ground truth.

**Supplemental Figure 3.**
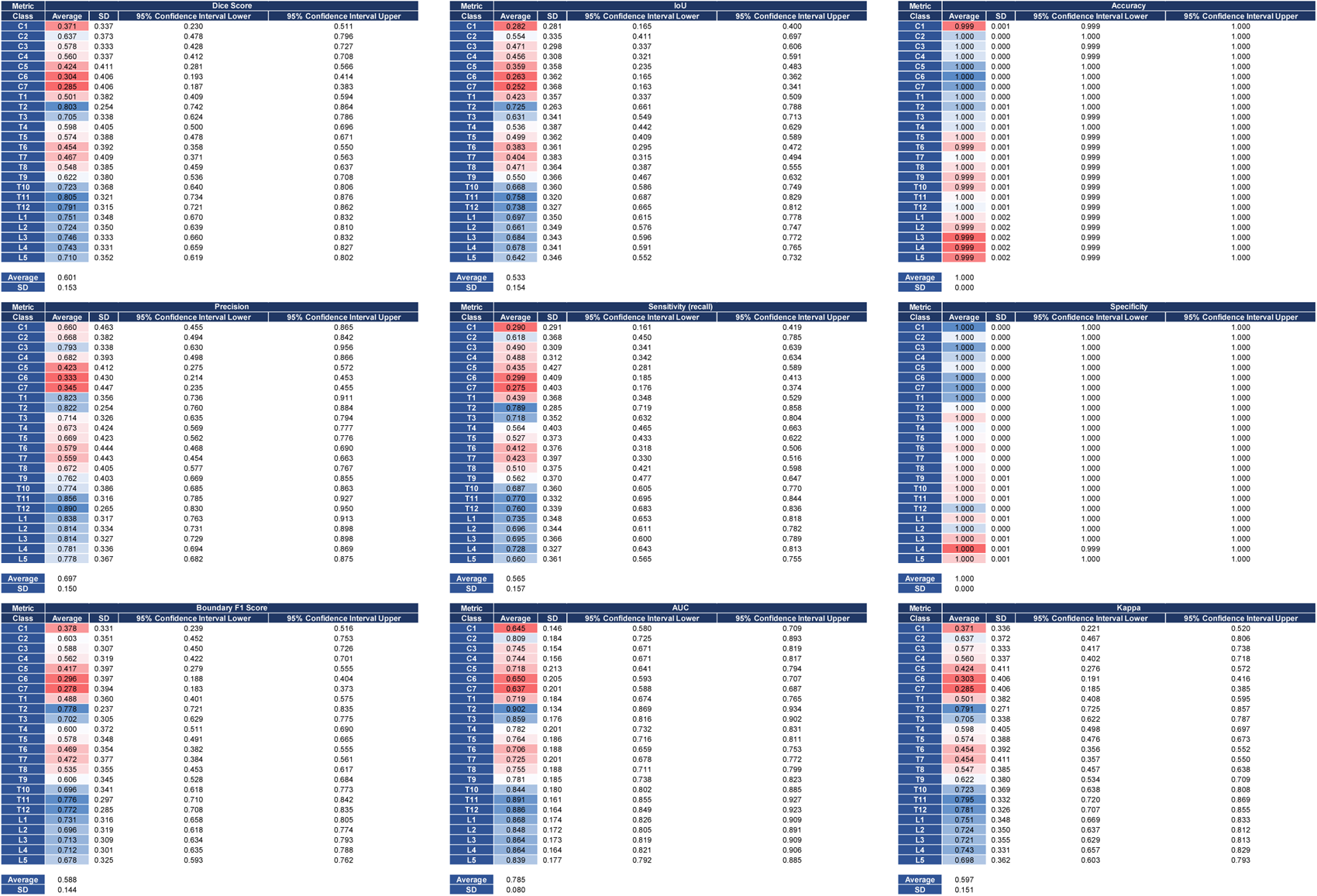
Comprehensive metrics evaluation of the open-source vertebrae segmentation model of MONAI-Label. Average metrics for the spineCT-ResSegNet model based on comparison between the predictions of the model on 108 datasets and their ground truth.

**Supplemental Figure 4.**
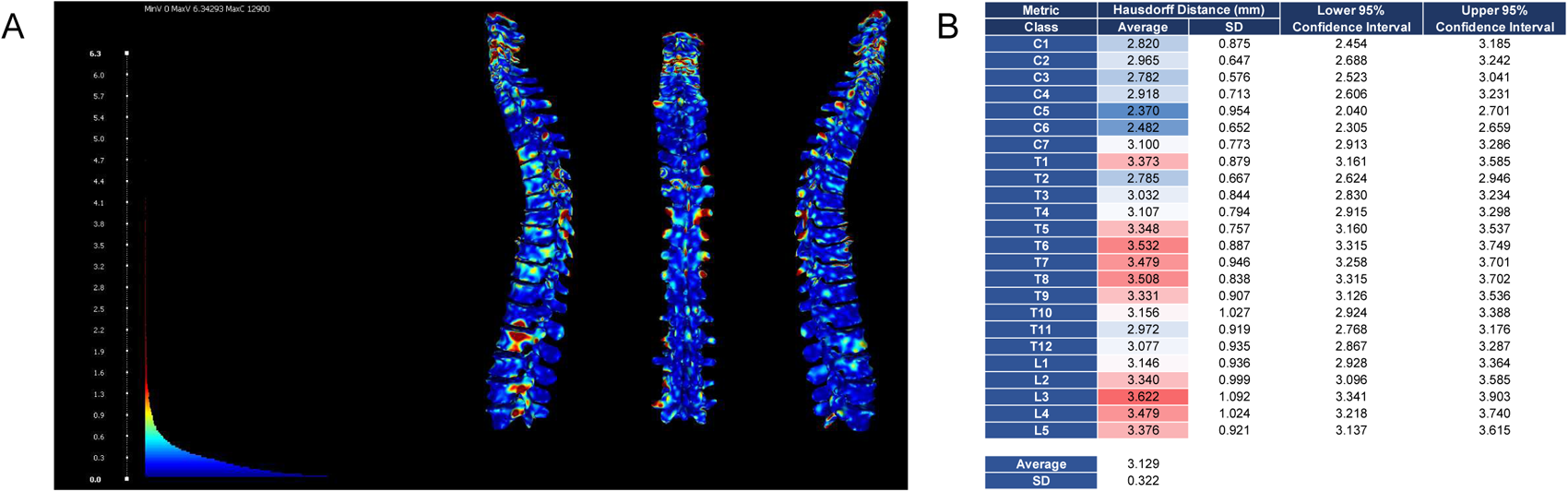
Hausdorff distance between prediction and ground truth by the open-source vertebrae segmentation model of MONAI-Label. (A) Example of the Hausdorff distance calculation between the spineCT-ResSegNet model (native to MONAI-Label) predictions and ground truth. (B) The pre-trained model present in MONAI-Label, trained on the VERSE dataset averages a Hausdorff Distance of 3.129±0.322 mm between model segmentation prediction and ground truth.

**Supplemental Figure 5.**
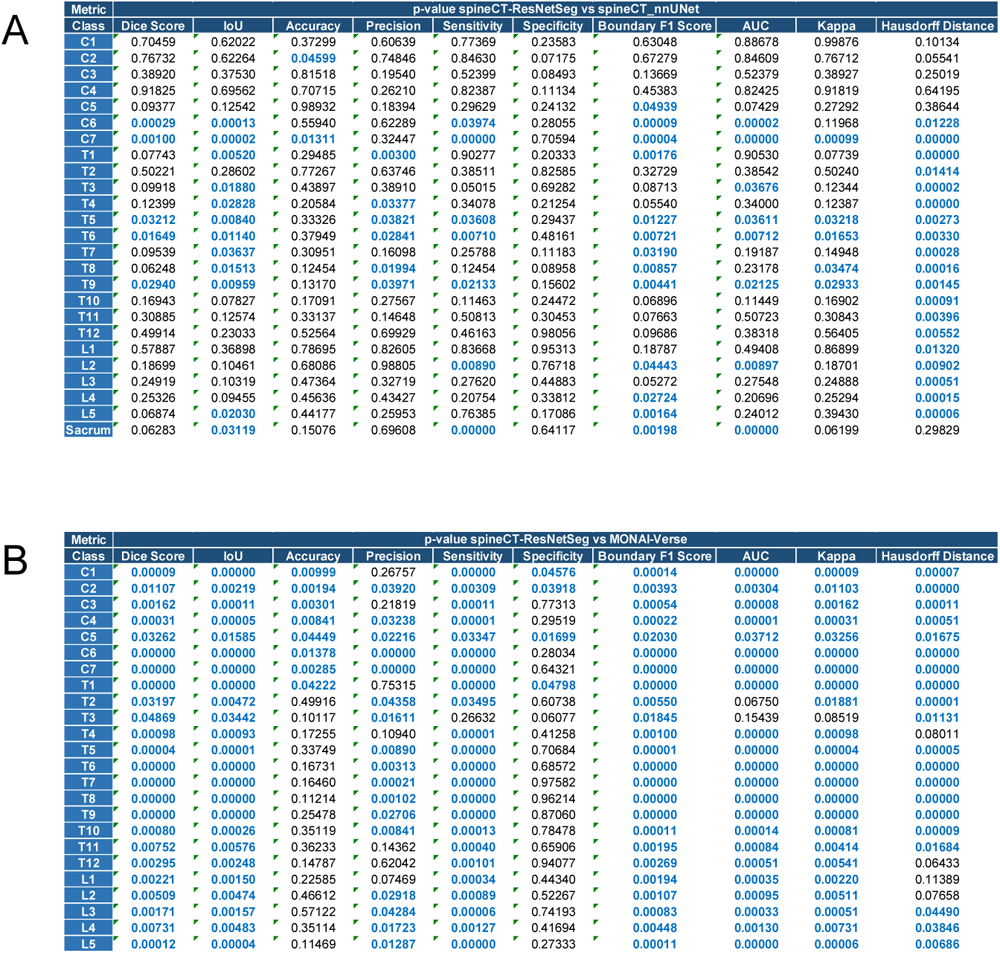
Statistical significance analysis between the quality segmentation metrics for each method used. (A) On average, both models, spineCT-ResSegNet and spineCT-nnUNet outputs are similar with the later one outperforming the MONAI-Label native model, only in regard to Hausdorff distance. Thus, both models appear to have a relatively equal prediction quality. (B) The newly trained spineCT-ResSegNet model mostly outperforms the pre-built spine segmentation model present in the MONAI-Label framework. Further, the model was trained in one extra class, the sacrum, capable of performing robust predictions for the anatomical class.

## Notes

### Competing Interest Statement

The authors have declared no competing interest.

### Funding Statement

Yes

### Author Declarations

N/A. All used data is open source and thus no special approvals were necessary.

